# Experimental efficacy of the face shield and the mask against emitted and potentially received particles

**DOI:** 10.1101/2020.11.23.20237149

**Authors:** Jean-Michel Wendling, Thibaut Fabacher, Philippe-Pierre Pébaÿ, Isabelle Cosperec, Michaël Rochoy

**Affiliations:** F-67000 Strasbourg, France, Occupational Health and Safety, Strasbourg, France.; F-67000 Strasbourg, Department of Public Health, GMRC, Strasbourg, France; NexGen Analytics, USA-82801 Sheridan, USA;, WY, USA; F-94210 St Maur-des-Fossés, France; Univ. Lille, F-59000 Lille, France; ULR 2694, CERIM, Public Health Department, Lille, France

**Keywords:** face shields, masks, particles, aerosolization, covid19, covid, coronavirus infections*, aerosols, pneumonia, viral*, prevention and control, protective devices, pandemics*, emitter, receiver, experimental setup

## Abstract

There is currently not sufficient evidence to support the effectiveness of face shields for source control. In order to evaluate the comparative barrier performance effect of face masks versus face shields, we used an aerosol generator and a particle counter to evaluate the performance of the various devices in comparable situations. We tested different configurations in an experimental setup with manikin heads wearing masks (surgical type I), face shields (22.5 cm high with overhang under the chin of 7 cm and circumference of 35 cm) on an emitter or a receiver manikin head, or both. The mannequins were face to face, 25 cm apart, with an intense particle emission (52.5 l/min) for 30 seconds. In our experimental conditions, when the receiver alone wore a protection, the face shield was more effective (reduction factor=54.8%), while reduction was lower with a mask (reduction factor=21.8%) (p=0.002). The wearing of a protective device by the emitter alone reduced the level of received particles by 96.8% for both the mask and face shield (p= NS). When both the emitter and receiver manikin heads wore a face shield, the ensuing double protection allowed for better results: 98% reduction for the face shields vs. 97.3% for the masks (p=0.01). Face shields offered an even better barrier effect than the mask against small inhaled particles (<0.3µm – 0.3 to 0.5µm – 0.5 to 1µm) in all configurations. Therefore, it would be interesting to include face shields as used in our study as part of strategies to safely significantly reduce transmission within the community setting.

## 1. Introduction

At the end of 2019, a novel coronavirus named Severe Acute Respiratory Syndrome Coronavirus 2 COVID (SARS-CoV-2) emerged [1]. The outbreak of the disease caused by this virus, Coronavirus Disease 2019 (COVID-19), was declared a pandemic by the World Health Organization (WHO) on March 11, 2020, and has caused nearly 1 million fatalities as of September 27, 2020 [2].

The airborne transmission route for SARS-CoV-2 is virulent for the spread of COVID-19 [3–5], as for SARS-CoV-1 [6]. At the present time, we have not identified the precise aerosol viral load or the minimum infectious dose of SARS-CoV-2 to cause an infection [7]. A viable virus can be emitted by an infected person by talking, singing, coughing or sneezing [8]: a small fraction of individuals are considered to be “speech super emitters”, releasing more particles than others [9].

A challenge in pandemic control is limiting the transmission of SARS-CoV-2 by asymptomatic or pre-symptomatic individuals [10]. A systematic review of the literature and meta-analysis revealed that face covering decreased the risk of airborne infections [11]. Surgical face masks significantly reduced detection of coronavirus RNA in aerosols [12]. Face covering by asymptomatic people (the primary case and family contacts before this primary case had symptoms) is effective in reducing transmission [13]. In the United States, an analysis revealed that the difference with and without mandated face covering represented the principal determinant in shaping the trends of the pandemic [4].

However, in Western countries, there has been significant controversy over the face covering [14,15], notably after the recommendation of the WHO on June 5, 2020 [16]. To improve compliance and acceptance, face shields are a good compromise and have many advantages: they are much more acceptable to young children [17], preferable during intensive aerobic physical activity, or for people who are anxious about wearing a mask [18]. Face shields could also be of economic and ecological interest because they are washable and therefore reusable.

There was controversy surrounding a recommendation to wear a face shield during this period, because little is known about the efficacy of different types of protective measures in the context of this pandemic [19]. CDC “does not currently recommend” the use of face shields as a substitute for masks because “there is currently not enough evidence to support the effectiveness of face shields for source control” [20].

It has already been shown that the use of face shields can significantly reduce healthcare workers short-term exposure to large infectious aerosol particles. Some authors think they are less effective against small particles (size less than 5 microns), which could remain suspended in the air for long periods of time and could easily get in through the wide holes on the sides and at the bottom and be inhaled. The space between the face and the face shield is indeed larger than that of the mask [21].

We hypothesized that face shields, worn by the emitter and worn by the receiver, could reduce the amount of particles from <0.3µm to 10µm emitted and received. We aimed to quantify the number of particles of different sizes detected in the mouth of a manikin head with different types of protections (face shield, mask or nothing); and to compare these different types of protection in different situations..

## 2. Materials and Methods

The evaluation was carried out in August 2020 on an experimental setup with two manikin heads positioned at 1.70m high and at 25 cm from each other (**Figure 1**). The tests were carried out in an empty closed room of 18.40m^2^ without drafts or mechanical ventilation, and with a sectional door of size 8.4m^2^ closed during the tests.

**Figure 1.**
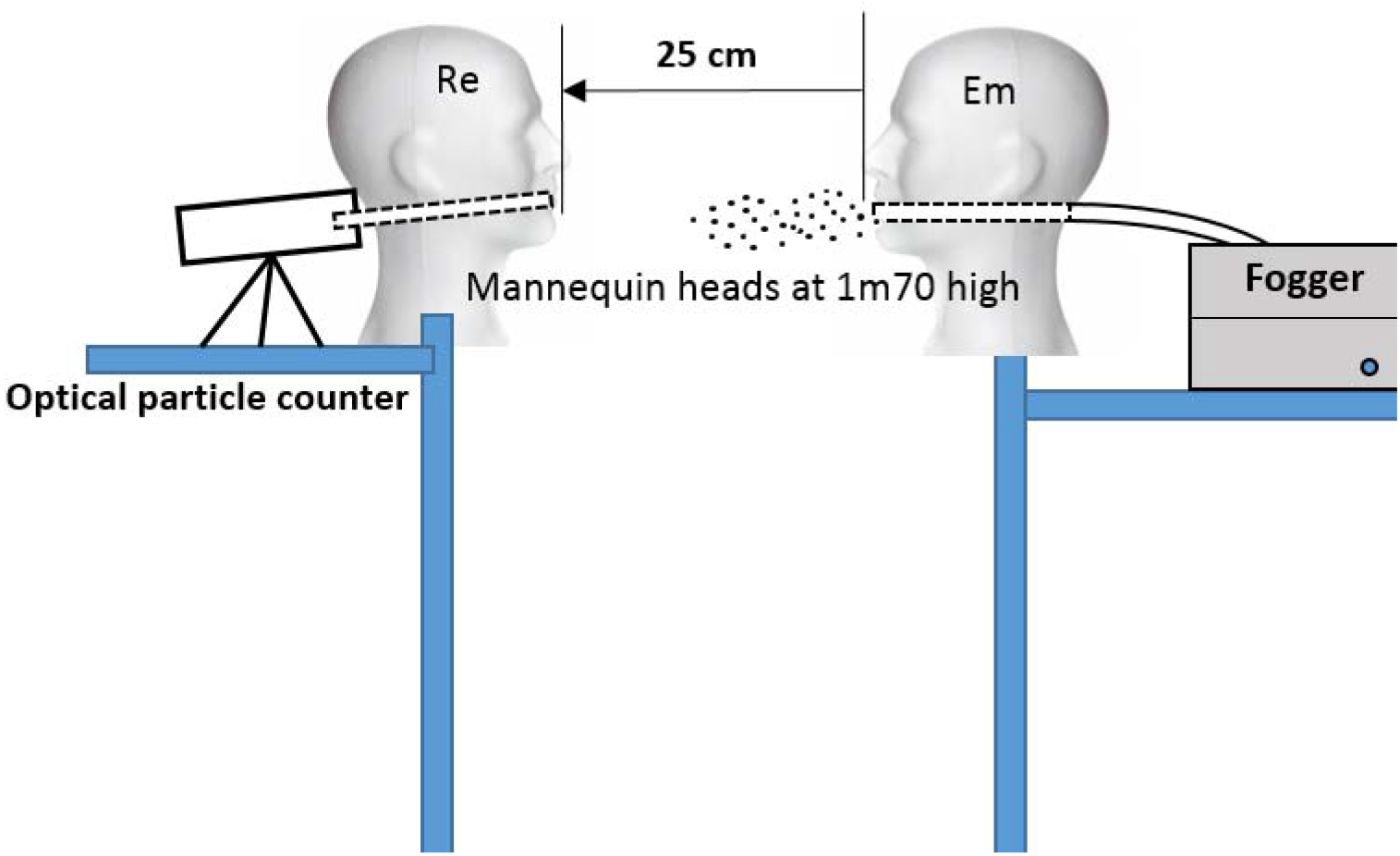
Experimental setup; Em: emitter; Re: receiver

One of the two heads (called an “emitter” or Em) has been hollowed out to reproduce a mouth. A pipe was introduced through a hole in the head to connect the fogger to the mouth. The atomizer was generated by aerosolizing distilled water with a fogger TRIXIE Fogger XL: the aerosol air flow coming out of the generator was continuous at 52.5 l/mn. The speed airflow mean value was 4.95 m/sec (sd=0.17m/sec) (n = 20).

The other head, called “receiver” (Re) has also been hollowed out at the mouth. A short pipe of 5cm long and 15mm internal diameter was connected to an optical particles counter.

The optical particles counter (TROTEC PC220) is designed to measure the size and the number of particles in the air. It sucks a volume of air for an adjustable amount of time and determines the size and amount of particles contained in it. The device is equipped with an integrated measuring cell with a laser (class 3R laser, 780nm, 1.5-3mW). According to ISO 21501, the counting efficiency is 50% at 0.3µm and 100% over 0.45µm.

Particles of sizes less than 0.3µm, 0.3µm to 0.5µm, 0.5µm to 1µm, 1µm to 2.5µm, 2.5µm to 5µm and 5µm to 10 µm were treated equally during the process. The cumulative counting method was performed for the analysis. The amount of all particles up to the selected particle sizes were counted (e.g.: “0.5µm = 417” means that 417 particles had a size between 0.3 and 0.5µm). The pumping time, air volume and the start delay are programmable. A HEPA filter was used on the counter to reset to zero before each measurement.

The temperature, ambient humidity and the aerosol flow velocity were recorded using a hot-wire thermo-anemometer TROTEC TA300. This device comes equipped with a hot-wire sensor and microprocessor technology for signal amplification. This combination guarantees precise measuring results.

Two personal protective equipment (PPE) were tested and their barrier performances were compared (**Figure 2**): EN14683 surgical masks type I (over 95% of 3µm particle filtration); face shields (Spanish brand) respectively: height covering the eyes, mouth, nose, 22.5 cm high with overhang under the chin of 7 cm, circumference of the visor 35 cm, front opening 4 cm high in line with the center of the forehead.

**Figure 2.**
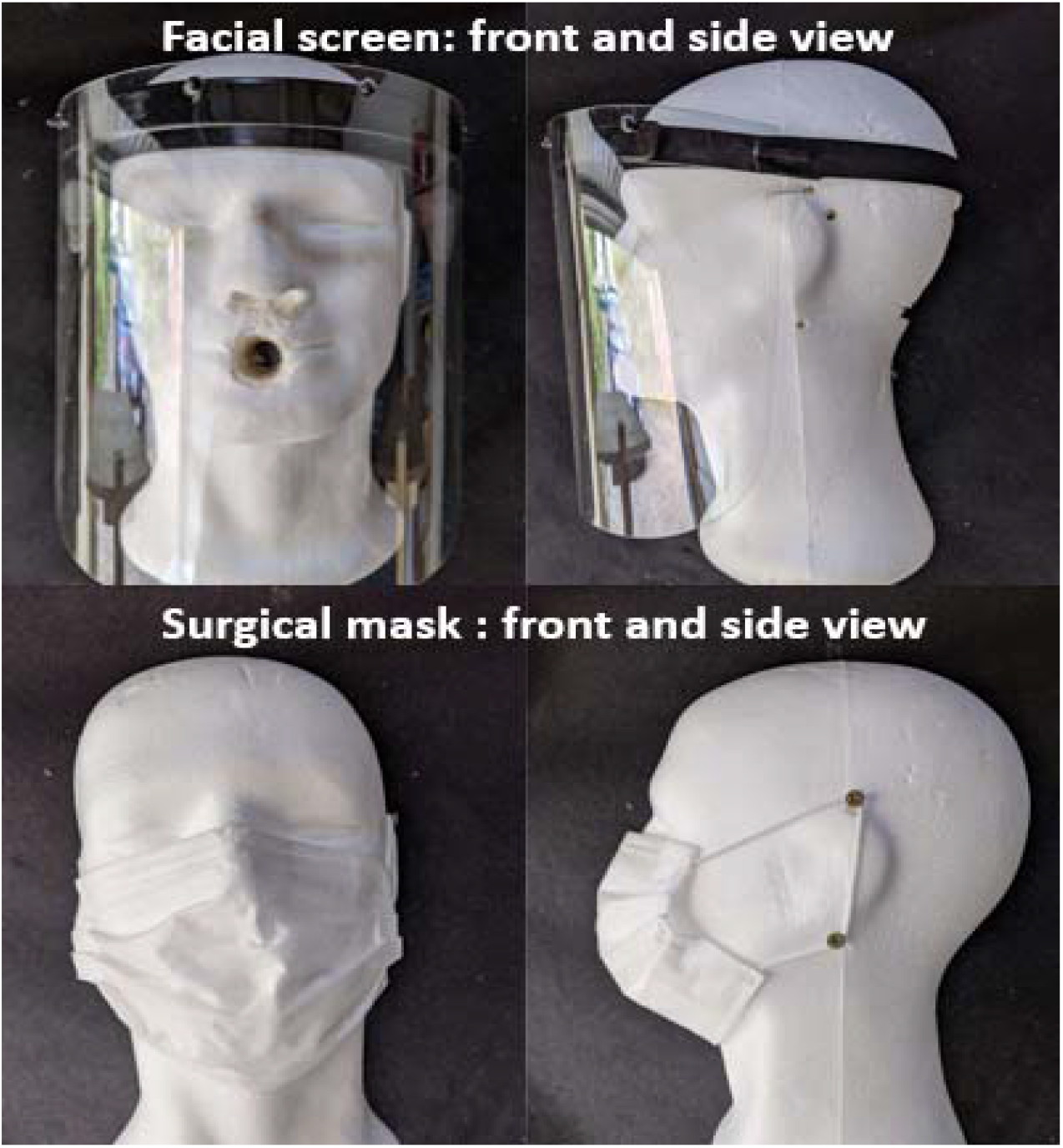
Surgical mask type I and face shield (front and profile views)

The background level of particulate pollution was first evaluated in the experimental room before (situation 0 with 10 measurements) and between each experiment (figure 3: 1 to 4a).

**Figure 3:**
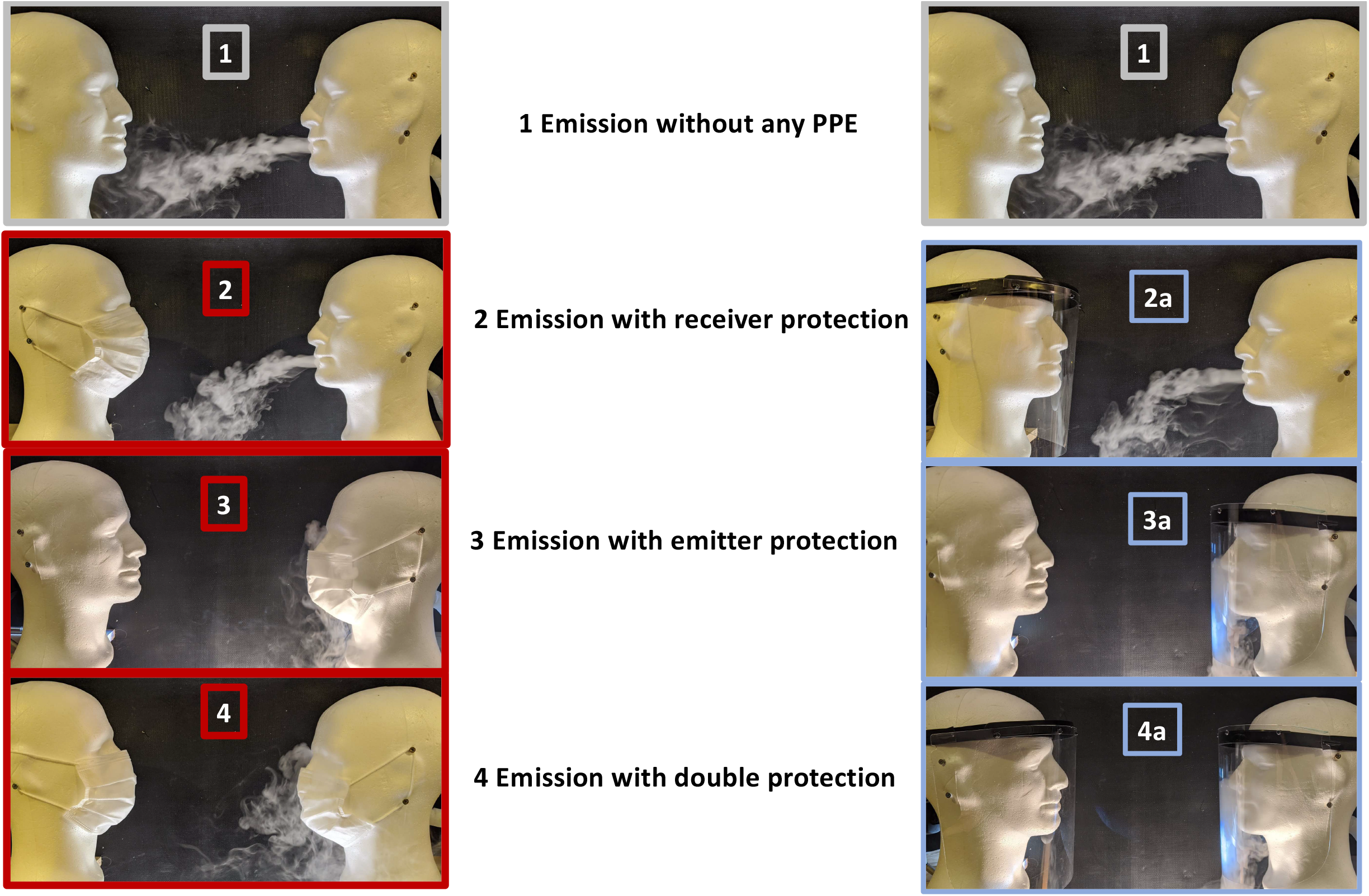
experimental configurations: 1 emission without any PPE - 2 emission with **Re**ceiver protection 3 emission with **Em**itter protection - 4 double protection **Em**itter and **Re**ceiver

The aerosol was then generated, without any PPE. The amount of particles and the size distribution of the aerosol were measured at a distance of 25 cm (10 measurements) (situation **1**). This configuration is the reference configuration with the maximum exposure of the receiver without any protection to which all the others will be compared.

Six configurations with PPE devices were then tested each time with a series of 10 measurements (**Figure 3**): surgical mask (situation **2**), then face shield (situation **2A**) on the receiver head only; surgical mask (situation **3**) then face shield (situation **3A**) on the emitter head only; surgical mask (situation **4**) then face shield (situation **4A**) on both emitter and receiver heads (double protection).

The aerosol generator was started with an airflow speed of 4.95 m/sec. The airflow speed of our generator was representative of the exhaled air velocity when talking [22]. The counter was started with a 5 second programmed delay from the generator, time to ensure that the head of the receiving manikin was well surrounded by the particle flow. The particle counter calculated the total cumulative particles aspirated on a volume of 1.416 liters. Counting was performed on the 6 channels (<0.3µm - 0.3 to 0.5µm - 0.5 to 1µm - 1 to 2.5µm - 2.5 to 5µm and 5 to 10µm) during 30 seconds of sucking in air. After each measurement, the counter was reset to zero by the HEPA filter and the room ventilated by opening the sectional door for at least 5 minutes to remove airborne particles. Before testing, the background particles level, temperature and humidity were recorded. Our Emitter-Receiver basic configuration was not changed during the entire experimentation: only masks and face shields have been exchanged for the data acquisition. First, the amount of particles and the distribution of the particle size at a distance of 25cm were studied without any PPE.

For the experiments performed with the aerosol particle measurement instruments, the parameters studied and compared were: the impact of the presence of PPE, the difference between protection of the emitter, receiver or both, the influence of each type of PPE, mask or face shield and the influence of the aerosol particle sizes on the results (<0.3µm - 0.3 to 0.5µm - 0.5 to 1µm - 1 to 2.5µm - 2.5 to 5µm and 5 to 10µm respectively).

We decided to evaluate and compare the barrier performance of each device in 6 different configurations by the Reduction Factor (**RF**) of the particles received and inhaled by the manikin head according to the following formula:

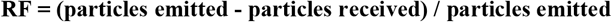

The percentage of inhaled particles (**PIP**) is obtained by the following formula:

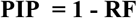

We also present the results of the barrier effect of each device (mask or face shield) using a particle size range approach.

For descriptive analysis we used mean value, median and standard deviation. To compare the reduction factors between the mask and face shield and the impact of location (emitter vs. receiver) of each protection, we performed a non parametric Mann-Whitney Wilcoxon test (with p-value computed for each comparison). We note that the independence assumption holds because the shield-group and the mask-group observations were successively measured in different runs of the same experimental setting. The alpha risk was set at 5% for all analyses.

## 3. Results

### 3.1. Environmental factors measurements

The temperature and hygrometry in the experimental room were regularly measured (n=20). The mean values were respectively 27.73°C (sd=0.50°C) and 68.3% (sd=2%). Before aerosolization, the thermo-anemometer confirmed the absence of significant air current in the room around the test bench (values < 0.05m/sec in all directions). During the production of aerosol by the fogger, in the first step of the study, the speed of the airflow coming out the pipe without any protection was evaluated at the mouth of the manikin head with the thermo-anemometer.

### 3.2. Aerosol total particle measurements

The background level of particulate pollution on the sucked in air volume standard in the experimental room showed a mean value of 13,837 particles (sd=2,436) (**Table 1**).

**Table 1.**
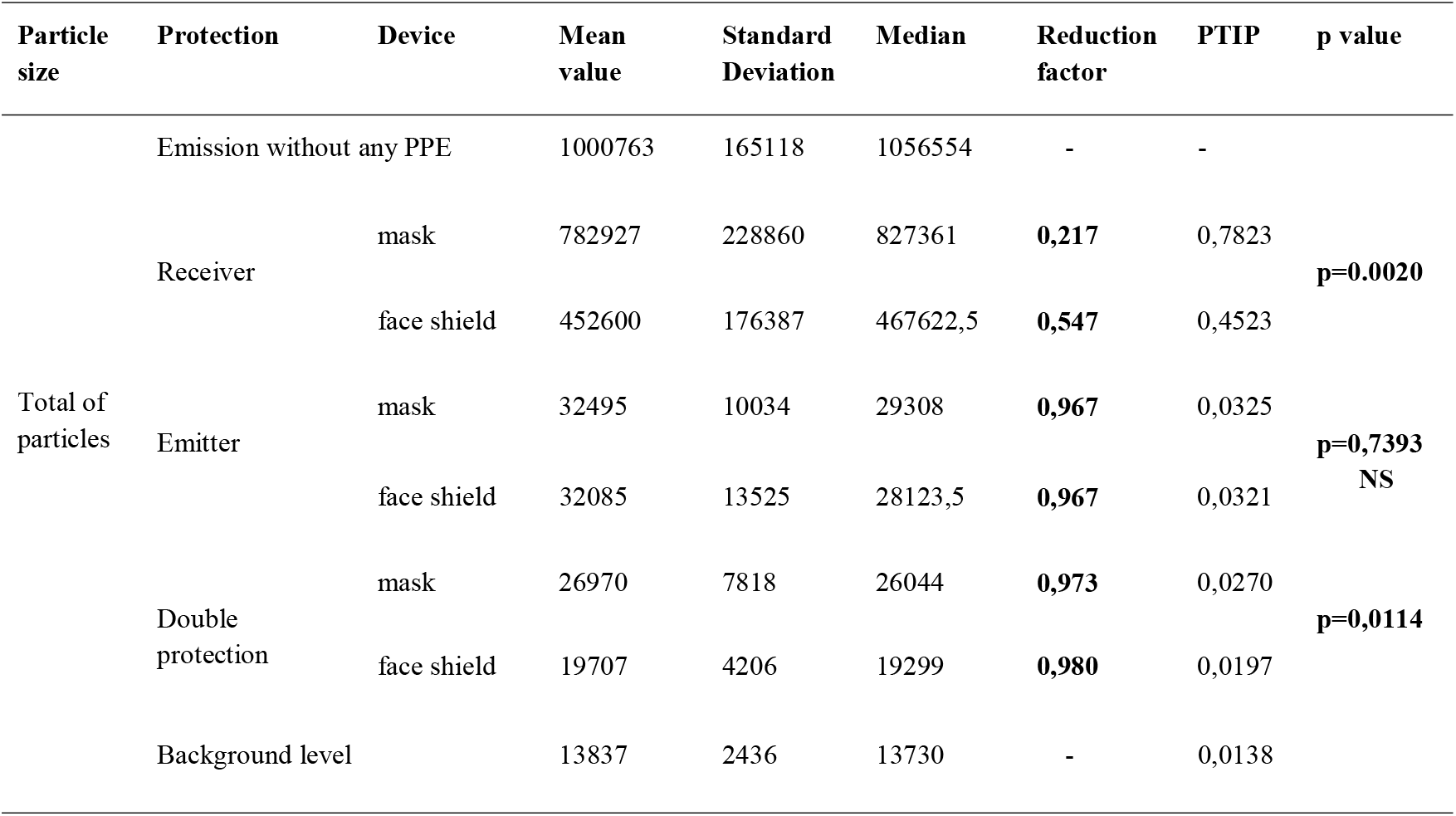
Reduction factor (RF) and Percentage of Total Inhaled Particles (PTIP) in the different configurations

The particles aerosol amount, received at 25 cm, decreased according to size. The average amount of particles measured at 25 cm without protection was 1,000,763 particles (sd=165,118). Of all the particles, 60% were less than .03µm and 94% less than 1µm (**Figure 4**). The number of detected particles decreased with increasing particle sizes.

**Figure 4:**
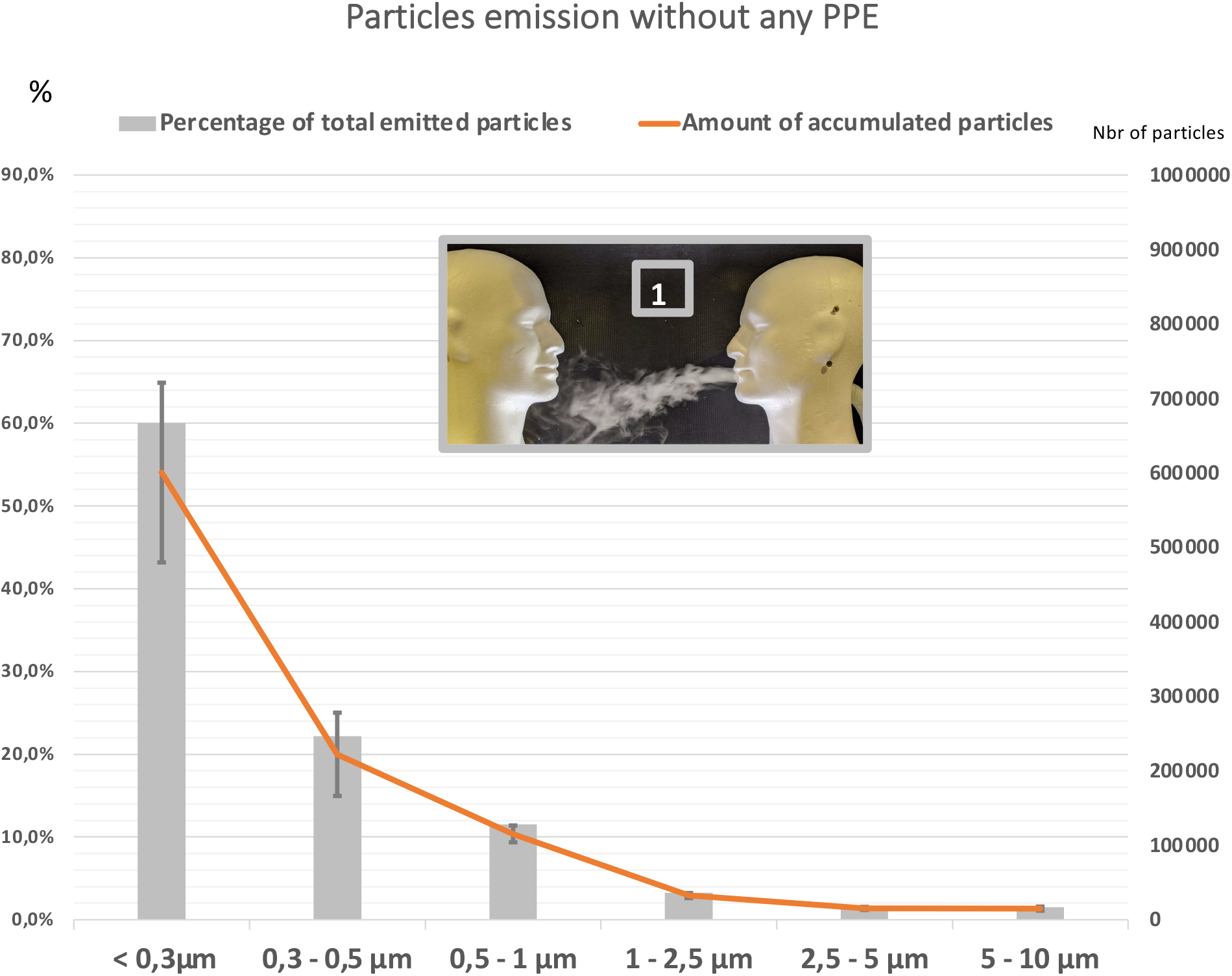
Particles distribution received at 25 cm without any PPE (situation 1)

### 3.3. Efficacy of face covering: total particle reduction factor depending on the situation

We evaluated the quantity of particles that were stopped by the different devices and calculated reduction factors (RF) and percentages of total inhaled articles (TPIP) in each of the 6 configurations. Thus the reduction factors were compared. When only the receiver head wore a mask, it significantly reduced the amount of total particles found by 21.8%; when the receiver head wore a face shield, the reduction was significantly higher: 54.8% (p = 0.002).

When the emitter head wore a mask or a face shield, it reduced the amount of total particles found by a greater level of 96.7% without any difference between the mask and face shield. When the receiver and the emitter wore a mask, it significantly reduced the amount of total particles inhaled by a greater level of 97.3%; when the two wore a face shield, the reduction was significantly higher: 98.0% (p = 0.0114) (**Table 1, Figure 5**).

**Figure 5:**
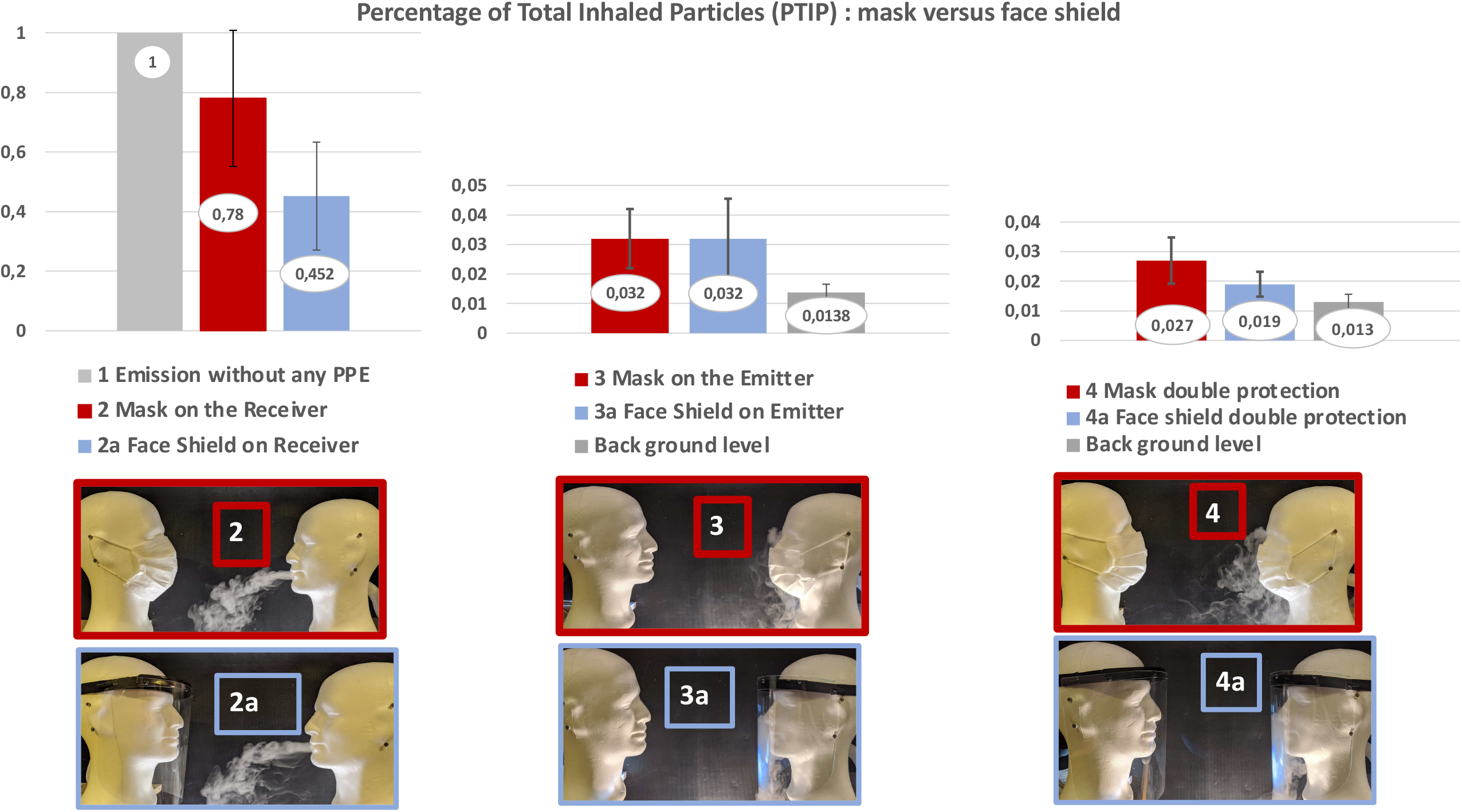
Reduction factors for the total particles: mask versus face shield

### 3.4. Efficacy of face covering depending on particle size range

Considering all particle sizes (≤0.3µm to 10µm) the reduction factors were always better or similar with the face shield compared to the mask. The face shield, when worn only by the receiver, was always more effective in blocking particles than the mask, and for all particle ranges. The face shield performed significantly better when the particles were smaller: reduction factors were respectively in range <0.3µm: 47.9% vs 13.8% for mask (p=0.006), in range 0.3-0.5µm: 47.6% vs 6.14% for mask (p=0.02), in range 0.5-1µm: 81.9% vs 60.9% for mask (p=0.009), in range 1-2.5µm: 92.3% vs 79.8% for mask (p=7.5.10^−5^) in range 2.5-5µm: 97.1% vs 91.9% for mask (p=0.009), in range 5-10µm: 99.6% vs 98.5% for mask (p=0.0006) (**Figure 6, Appendix table 1**).

**Figure 6:**
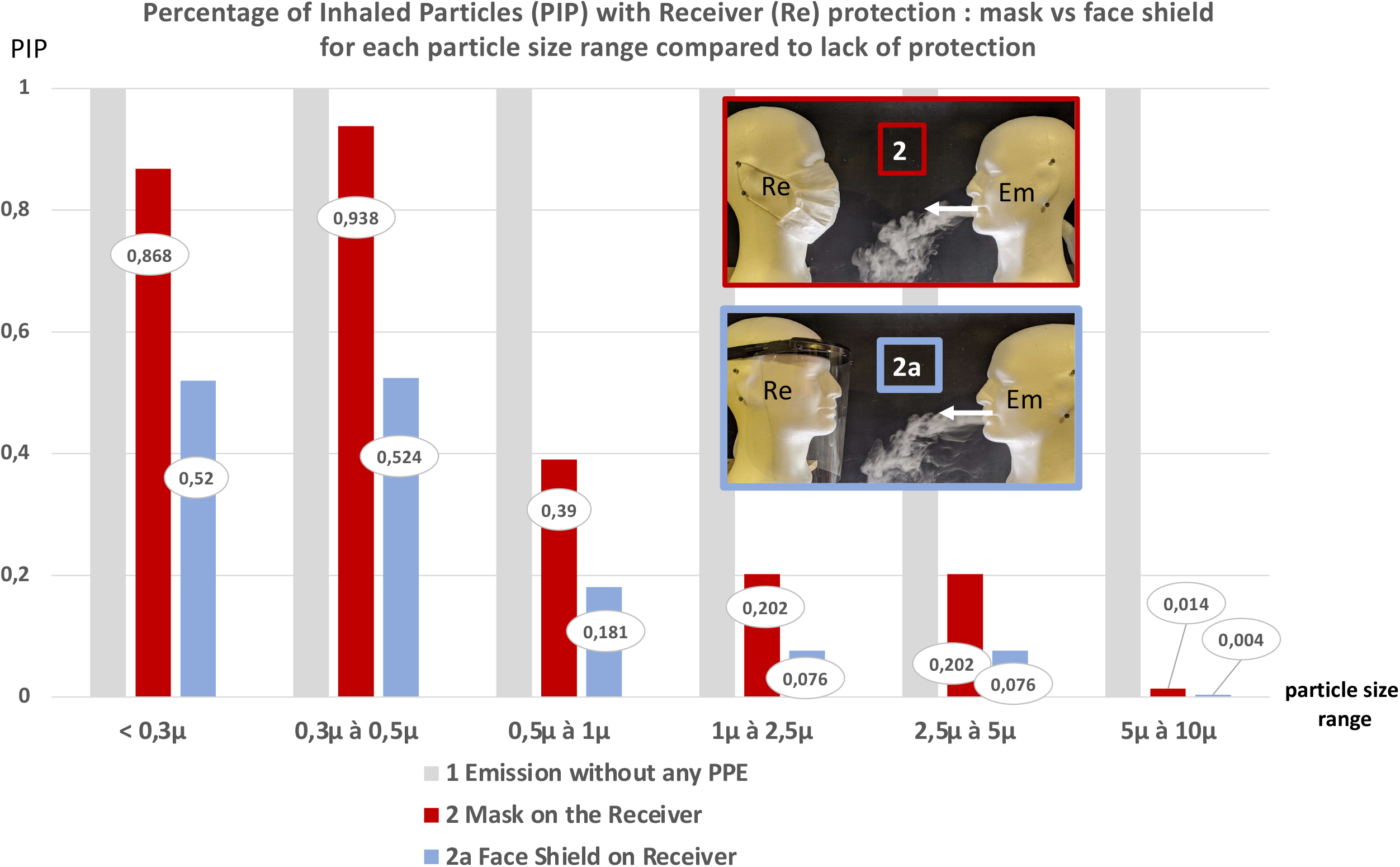
Percentage of Inhaled Particles (PIP) with Receiver (Re) protection: mask vs face shield for each particle size range compared to lack of protection

The face shield, when worn only by the emitter, performed about the same as the mask for all sizes of particles with more than 96% reduction factor for the particles less than 1µm and 99% reduction factor for the particles more than 1µm in size (**Figure 7, Appendix Table 1**).

**Figure 7:**
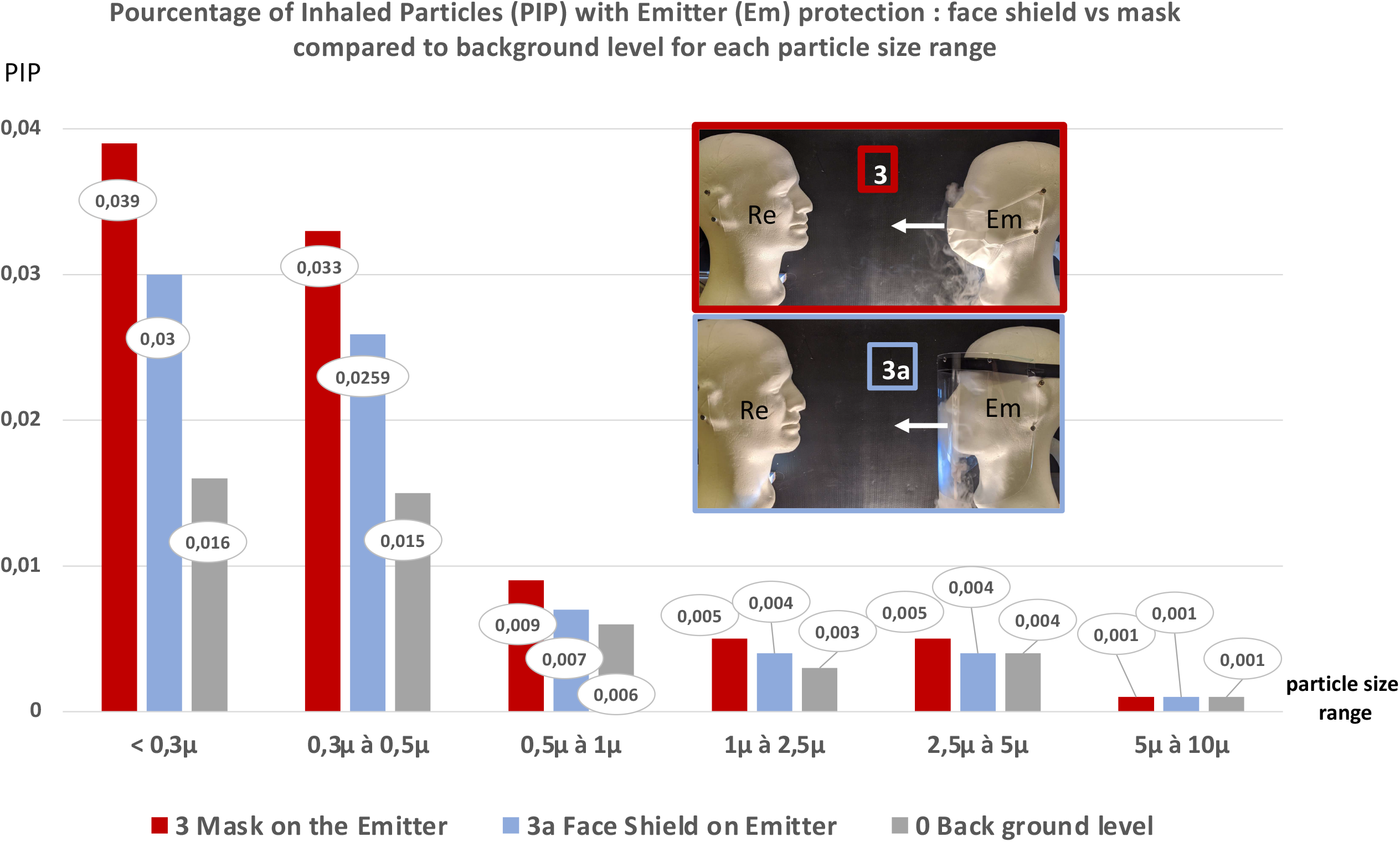
Percentage of Inhaled Particles (PIP) with Emitter (Em) protection: mask vs face shield for each particle size range compared to lack of protection

When double protection was worn, the face shield performed significantly better with a reduction factor of 97.7% vs 96.8% for the mask in the range of < 0.3µm (p=0.01), 97.8% vs 97.1% for the mask in the range 0.3-0.5µm (p=0.052 NS). For particles more than 1µm in size, masks and face shields worn by the emitter and the receiver reduced the amount of inhaled particles by more than 99% (**Figure 8, Appendix Table 3**).

**Figure 8:**
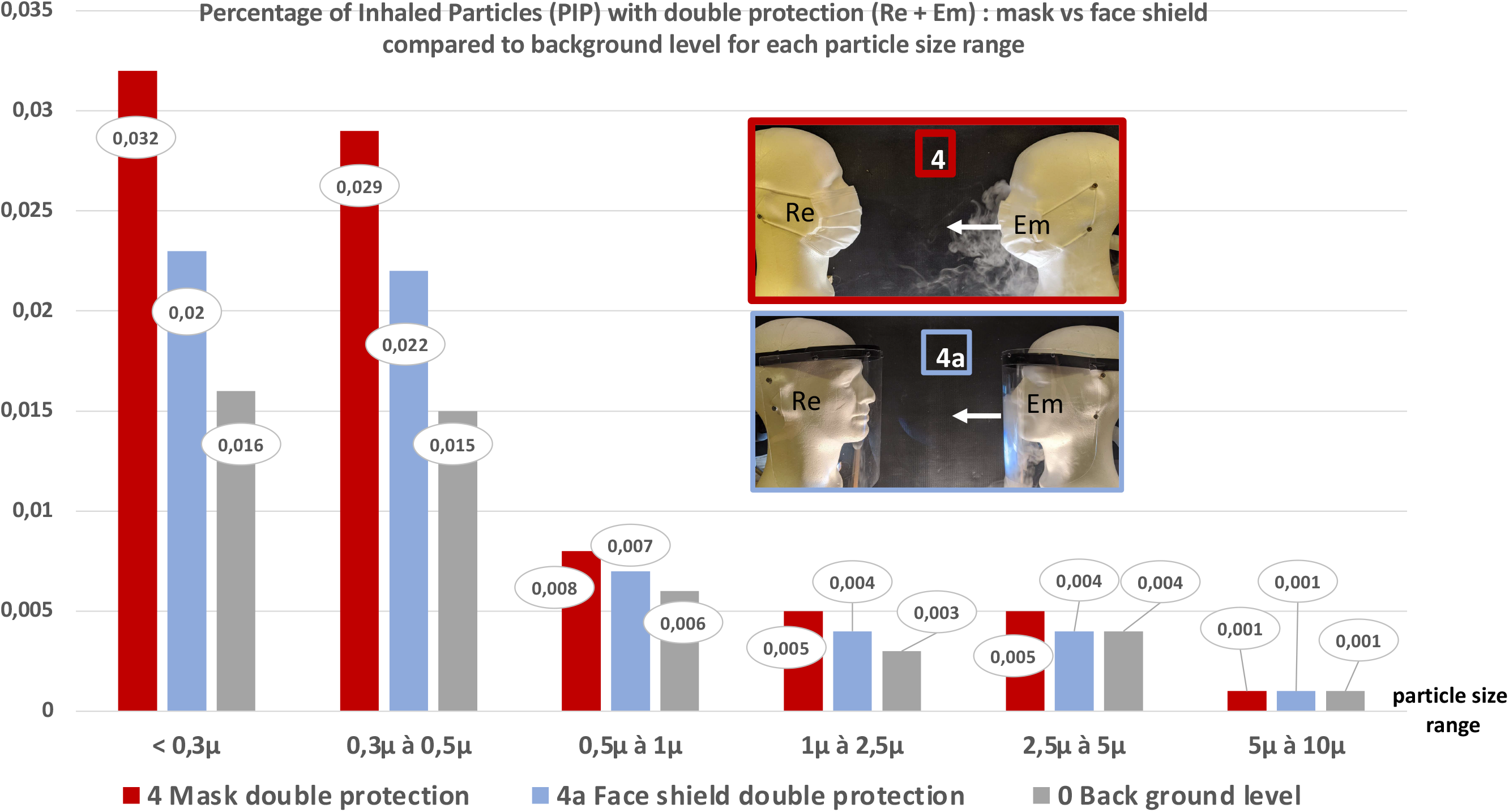
Percentage of Inhaled Particles (PIP) with Double protection (Em+Re): mask vs face shield for each particle size range compared to lack of protection.

### 3.5. Comparison between emitter or receiver protection

Statistically fewer particles are detected when the protection is worn by the emitter alone (RF=96,7%) compared to the protection worn by the receiver alone (p=10^−5^) (**Table 2**).

**Table 2.** Reduction factor (RF) and Percentage of Total Inhaled Particles (PTIP) relating to the location of the protection

|  | Location | Mean value | Standard deviation | Median | Reduction factor | PTIP | p value |
| --- | --- | --- | --- | --- | --- | --- | --- |
| **Mask** | Receiver | 782927 | 228860 | 827361 | **0,217** | 0,782 | $p=10^{-5}$ |
|  | Emitter | 32495 | 10034 | 29308 | **0,967** | 0,032 |  |
| **Face shield** | Receiver | 452600 | 176387 | 467622 | **0,547** | 0,452 | $p=10^{-5}$ |
|  | Emitter | 32085 | 13525 | 28123 | **0,967** | 0,032 |  |

## 4. Discussion

Few studies have evaluated the benefits of face shields in limiting infectious transmission (speaking, singing, sneezing, coughing, etc.) when worn by infected persons, whether symptomatic or not. In our experimental situation (short but intense exposition in a face-to-face situation), face shields and masks could reduce the amount of particles emitted and finally received by the target. The number of inhaled particles detected on the receiver decreased mainly when the emitter wore face protection, and more so when both the emitter and receiver wore face protection. Our results indicate a better outcome when the emitter wears face protection.

CDC currently considers that the main route of contamination would be by close contact and not by aerosolization at distance [5]. As part of the fight against the spread of SARS-CoV-2, face masks have been generalized to prevent propagation from asymptomatic emitters, as effective “anti-droplet screens” [23–25]. Another at-risk situation for SARS-CoV-2 transmission may be a manuporting route: indeed, SARS-CoV-2 can survive up to 9 hours on human skin, much longer than influenza-virus, and is found in wastewater and stools [26–28]. Surgical masks may be less protective for this pathway than face shields that prevent finger-to-eye contamination in addition to hand-to-mouth; however, current evidence does not suggest that this manuporting pathway is predominant [5]. The current hypothesis is that particles emitted when speaking or breathing are mainly formed by a “fluid film bursting” mechanism inside the small airways of the lungs and/or by the vibration of vocal folds in the larynx [4,29]. The amount of aerosols emitted during speech correlates with the loudness of vocalization, physiological factors and language spoken, ranging from approximately 1 to 50 particles per second, with some “super emitters” which release much more particles in quantity than their peers. Particle emission when speaking shows a pic size of 0.75 to 1µm and maximum size around 7µm [9,22,30,31]. The particles smaller than 1µm were easily reproduced by our aerosol generator.

Under our experimental conditions, the total number of particles received was significantly lower when wearing a face shield than when wearing a mask by the receiver, especially for micro-particles around and less than 1µm, the ones which are emitted during speaking. Face shields are commonly used by health workers to protect the face; CDC strongly recommends to wear a face shield covering the front and sides of the face, a mask with an attached shield, or a mask and goggles, during aerosol-generating procedures on patients not infected with M. tuberculosis, SARS, hemorrhagic fever viruses or other viruses requiring the use of N95 type protections [32].

Lindsley and al. studied face shields and concluded that they can reduce healthcare workers short-term exposure to large particles of infectious aerosols. The effectiveness of the face shield, reducing the amount of inhalation exposure to influenza virus on the receiver was estimated to range between 68% and 96%. After 30 minutes, the effectiveness was 80% and face shields stopped 68% of aerosols [21]. One recent preprint study found that the face shield blocked more than 90% of the otherwise inhaled particles; for finer particles (0.3μm), the face shield performed much better, as in our study [33].

If a transmitter wears a mask or face shield, the source control of inter-individual transmission is expected to improve [18,34]. Surgical masks are now the gold standard in the fight against COVID-19 to control the source emission. There are some limitations with the surgical mask, including potential permeability to particles less than 3 microns, leaks even if the filtration performance is announced at more than 95%, often improper wear and variable acceptability. Despite these limitations, its effectiveness has been widely demonstrated [12,14,16,20,23,24,35,36]. To our knowledge, few studies have compared the ability of source control between face shields and masks in an identical configuration or in a dual protection (emitter and receiver). Face shields were often used according to the paradigm of personal protection, as before the pandemic in Western countries [15].

Our results differ from those of Verma et al.: to evaluate the performance of the face shield as source control, they used a cough simulator, synthetic smoke and two lasers (horizontal and vertical); by placing a plastic face shield they found that smoke particles spread behind the emitter [37]. However, they did not quantify the number and the distribution of particles emitted, or the decreased concentration with distance. In addition, they did not use an aerosol consisting of water-based liquid particles, but a smoke generated at high temperature, which behaves differently. The face screen was positioned semi-open, in an improper way, facilitating the exit of a plume of exhaled air; in contrast to Verma et al., ve used a liquid aerosol generator mimicking the particles emitted by the voice at room temperature, with a right angle position close to the chin down, a chin and forehead overhang, and most importantly, a position of the visor parallel to the face [37].

Some other experimental studies used collection chambers with a manikin head placed in direct contact with the walls and in very small enclosed spaces, sometimes with an air suction device [38, 39]. Our experimental conditions are closer to a real-life situation: we take into account the effect of air dilution in an open space with heads located at the height of standing, average sized individuals. However, these results are also in line with those showing that mask protection is never completely effective in small, poorly ventilated enclosed spaces; moreover, the wearing of facial protection must be always associated with respect for physical distancing [36, 40].

In another experimental pre-print study [41], researchers used Background Oriented Schlieren (BOS), imaging to compare several types of face protections (surgical mask, homemade mask, commercial face shield and a 3D printed face shield). The results showed that during quiet, heavy breathing or coughing, no front throughflow was discernible for the lightweight printed 3D face shield. For surgical masks, exhaled air travelled more than 23.7 cm. The maximum distances travelled by the air flows (crown, brow, side and back jets) were all greater for the surgical mask compared to the face shield. Consistent with our observations, face shields generate particularly strong air flows towards the ground while no flow was discernible with the surgical mask in that direction: of course, it seems more interesting to direct the flow of infected particles towards the ground rather than letting them escape in the direction of the receiver.

Arumurua and al. compare the “barrier” performances against saliva spray emitted on different devices using a camera and laser illumination. During a sneeze projecting aerosols at 25 feet, the distance was respectively reduced to 2.5 feet with the polypropylene surgical mask (−90%), 1.5 feet with the double layer cotton mask (−94%), 1.5 feet with the triple layer cotton mask (−94%) and only 1 foot with a polycarbonate face shield (−96%). The study shows that a face shield had the best performance of all the devices and that the surgical mask is the one that provides the weakest barrier to sneezing. With an N-95 mask, the flow has been visualized at a distance of 2 feet behind the manikin [42]. So, face shields alone do seem to be a good alternative to protecting others. The generalized use of face shields should be considered to protect the environment, since experimental efficacy has been shown and found in those studies.

Face shields are commonly used as an infection control alternative to goggles, because they also protect other areas of the face (forehead, preauricular area, cheeks, chin, etc.) and limit splashes from the face especially to the eyes. A significant parameter is that face shields reduce the potential for contamination through the eyes. This property is particularly interesting in the fight against COVID-19. This route is rarely studied and the eye protection could be determinant in community settings [11,43]. A study performed in Hubei hospital found that only 5.8% of COVID-19 patients (16 of 276 patients) wore glasses compared to an estimated 31.5% of the general population, suggesting that eye protection, eyeglasses or face shields could be useful against COVID-19 [43]. The ocular mucosa and the nasopharynx are connected by the nasolacrimal duct. When splashes reach the cornea or conjunctiva, they can penetrate the nasolacrimal duct and be transported to the nasopharynx and trachea [44,45]. Ocular manifestations seem frequent, and high□frequency hand-eye contact correlates with conjunctival congestion [46,47]. Immunohistochemical analysis also revealed the expression of ACE2 and TMPRSS2 in the conjunctiva, the limbus and the cornea, which suggests that cells on the ocular surface are susceptible to SARS-CoV-2 and could therefore serve as a gateway and reservoir for interindividual transmission [44,48,49]. In a meta-analysis, eye protection was associated with a lower risk of infection in 13 studies (RR= 0.34) and 2 adjusted studies (aOR=0.22). Eye protection alone reduced the infection risk by 78% (aOR = 0.22) [11].

In addition to eye protection, face shields offer a number of advantages [18,19]. They are easily reusable and washable with soap or antiseptic [50]. The economic interest is obvious for the poorest populations throughout the world. They prevent the wearer from touching their mouth and nose. The face shield does not need to be constantly readjusted like the mask. The face shield, if its external surface is contaminated by the hands, can not be a source of external contamination, unlike the mask, because the screen is watertight. The face shield is better tolerated during intense physical effort. It prevents the appearance of mist on the corrective glasses and thus reduces the risk of accidents that occur while driving. They should be better tolerated in children, for whom the importance of facial protection is increasingly demonstrated in the fight against the population spread of COVID-19 [51]. People who wear a medical mask sometimes take it off to communicate, for comfort or to facilitate lip-reading, especially when dealing with people who are hard of hearing [52]. The face shield makes it easy to visualize important facial expressions in emotion readings, which is a crucial part in communication.

However, our study has some limitations: the experimentation configuration is an extreme configuration of exposure. Considering human particles (p) emission levels (breathing = 0.31 p/s, speaking = 2.77p/sec, coughing = 10.1 p/s) [35], our aerosol generator produced much more particles with a very short distance and short time of 30 seconds in an enclosed, unventilated space. Particles inhalation by aerosolization could also be achieved by long, low-intensity exposure: this is not tested here in our study. The study was only conducted on one type of surgical mask type I and one type of face shield model. Real life leads to situations where the emitter is not always facing the receiver, but we made the ballistic assumption that the riskiest situation was face-to-face to compare surgical masks and face shields in the exact same configurations [9,30]. The way in which one wears a mask or face shield, in addition to the shape and design of these protections, can be a determining factor in the efficiency of its performance [41]. Further experimentations should confirm performance through the shape and the face shield design. To protect others in the best possible way, the objective should be to send the exhaled air flow towards the wearer or towards the ground with the most efficient design of the visor. More studies are needed in real-life situations to determine the best possible uses and effectiveness of face shields. Some microbiological studies on sick people equipped with face shields with environmental surfaces analysis and on air bio collectors should be conducted.

## 5. Conclusions

Our results show that, in our experimental conditions, when the receiver alone wore a face shield, the amount of particles was significantly reduced compared to when the receiver alone wore a mask, even with small particle size emission (≤0.3µm). When the emitter wore a face shield or mask, it was more efficient than when only the receiver wore protection, with no difference between mask or face shield. In our conditions, the double protection allowed for even better results: 98% reduction for face shields and 97.3% for masks. Furthermore, face shields are durable, can be reused and are easy to clean. They prevent the wearer from touching their mouth, nose and especially eyes, a possible route for the transmission of SARS-CoV-2. The efficiency of the face shields also encourages the development of their use where no facial protection is possible (school canteens, bars…), through the integration of Plexiglas between users. Finally, the results of our experimental study suggest that face shield should be included as part of an expanded arsenal against SARS-CoV-2, as a possible face covering alternative for people (in priority children, disabilities or medical intolerant to masks), for situations where no facial protection is used (collective sporting or cultural practices without masks) or in addition to masks.

## Data Availability

Data are available in manuscript.

## Supplementary Materials

The following are available online at www.mdpi.com/xxx/s1, Figure S1: title, Table S1: title, Video S1: title.

## Author Contributions

Conceptualization, JMW, MR; methodology, JMW, TF, PPP; software, JMW, PPP; validation, TF, JMW, MR; formal analysis, PPB, TF; investigation, JMW; resources, JMW; data curation, TF; writing—original draft preparation, MR, JMW, TF, IC; writing—review and editing, MR, JMW, PPP, TF, IC; visualization, JMW, TF, MR; supervision, JMW, MR. All authors have read and agreed to the published version of the manuscript.

## Funding

This research received no external funding.

## Acknowledgments

No.

## Conflicts of Interest

The authors declare no conflict of interest.

## Appendix

**Table 1.**
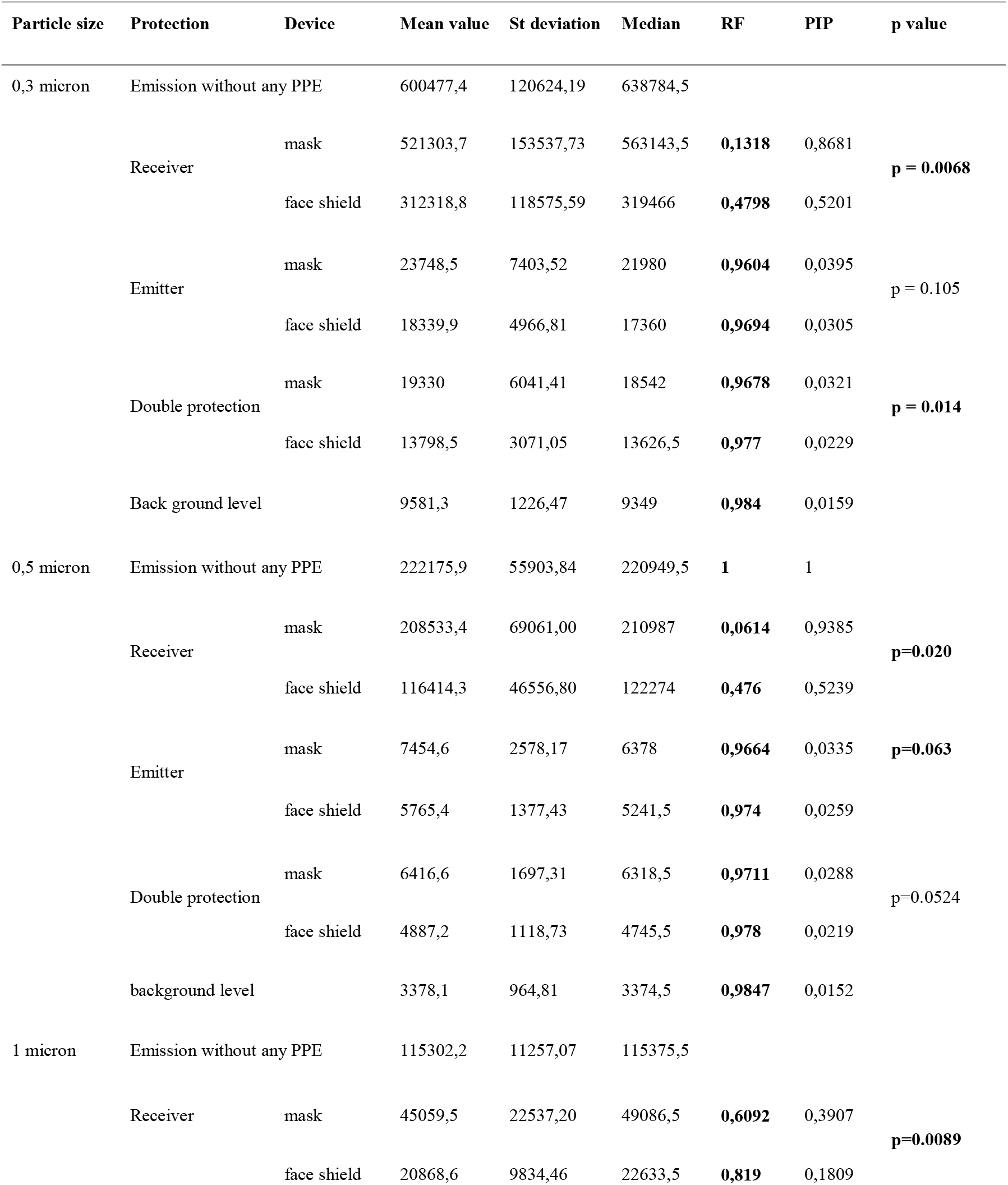
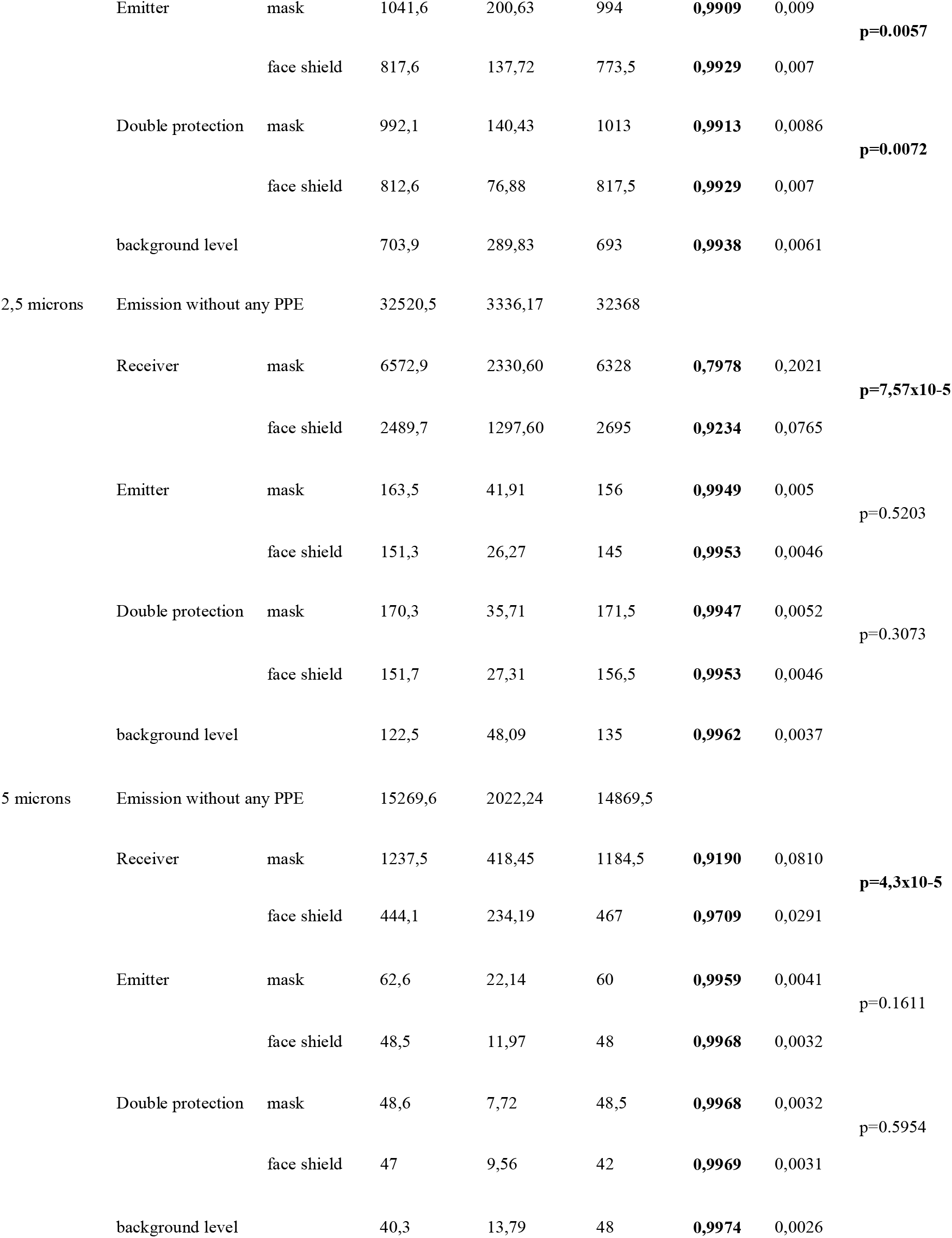
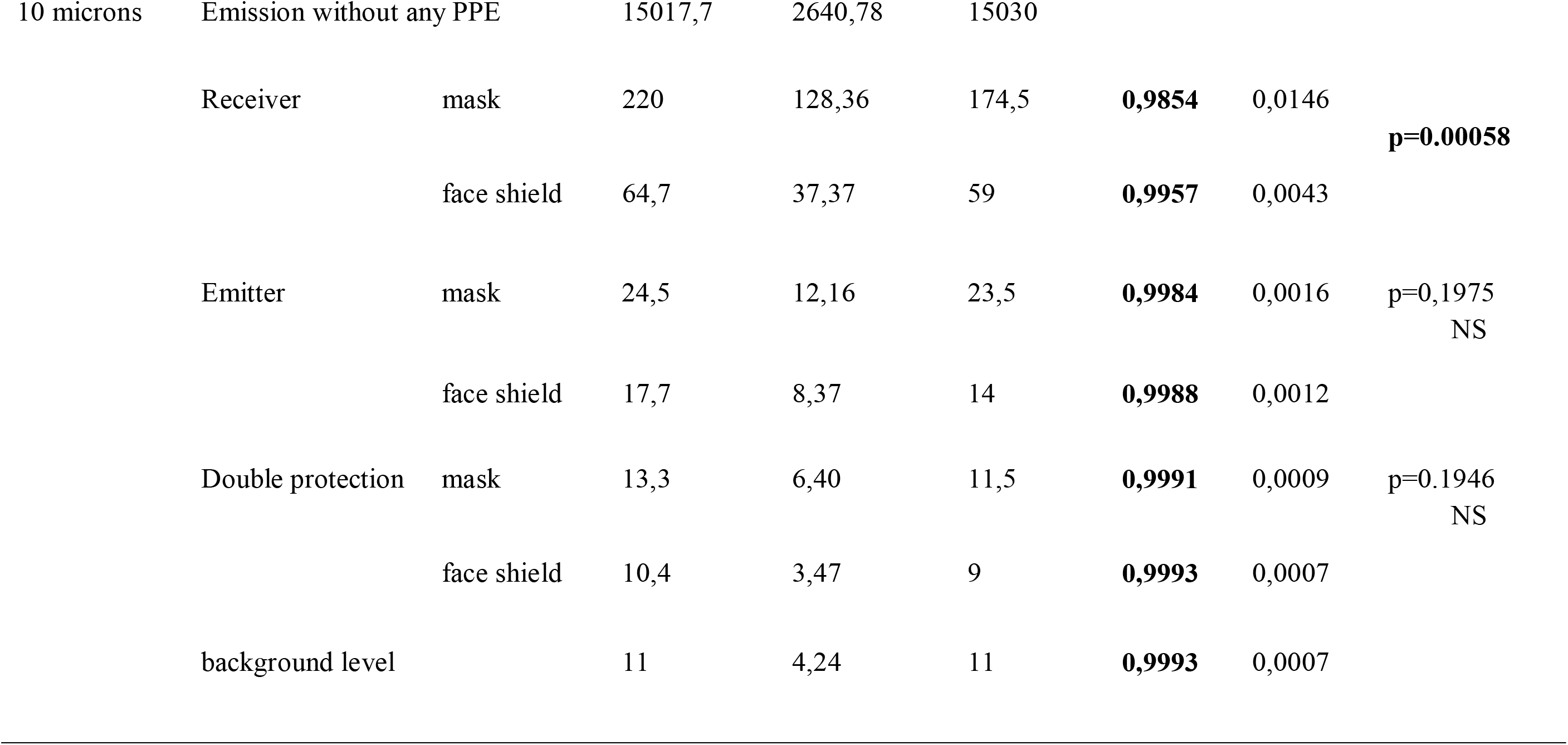
Results of aerosol particle measurements on each particle size range in the different configurations

## Notes

### Competing Interest Statement

The authors have declared no competing interest.

### Funding Statement

No external funding was received

### Summary of Updates

Revision of the text (without changing the results) with clarification and new graphics, at the request of the journal being proofread.

