## Appendix Figure 1 for "Experimental efficacy of the face shield and the mask against emitted and potentially received particles" (from another version of this preprint: version 1, 2020-11-24)

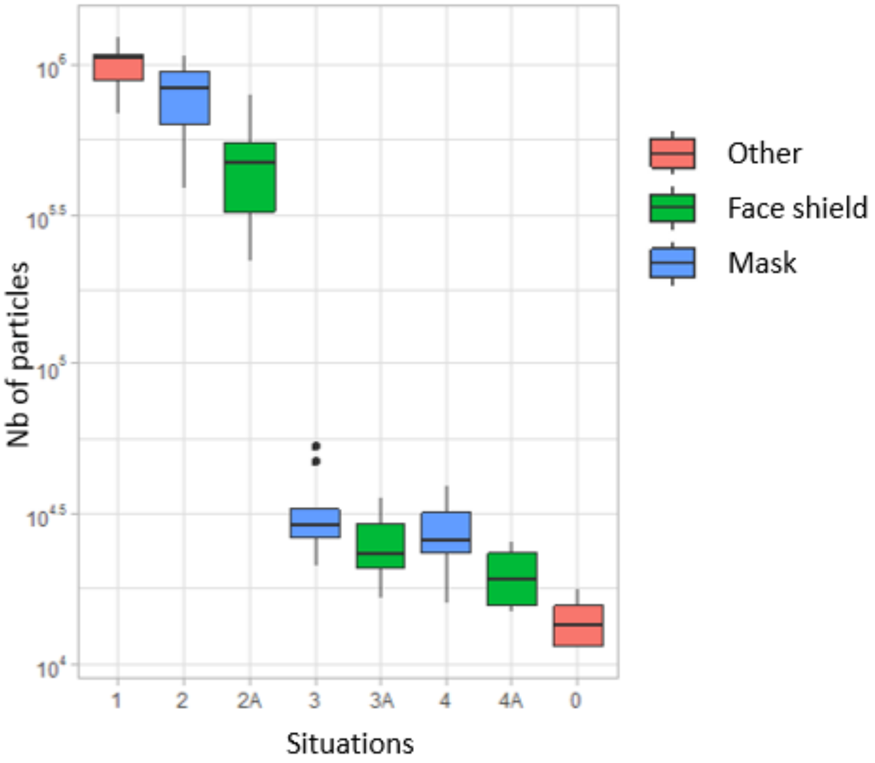


**Figure 1:** Aerosol emission at 25 cm with or without PPE

Situation 1: Without protection; Situation 2: Mask on receiver; Situation 2A: Face shield on receiver; Situation 3: Mask on emitter; Situation 3A: Face shield on emitter; Situation 4: Mask on receiver and emitter; Situation 4A: Face shield on receiver and emitter; Situation 0: background noise.
