## Appendix Table 1 for "Experimental efficacy of the face shield and the mask against emitted and potentially received particles" (from another version of this preprint: version 1, 2020-11-24)

**Table 1:** Results of aerosol particle measurements

| Experience | Median (N) | Min (N) | Max (N) | First quartile (N) | Last quartile (N) |
|---|---|---|---|---|---|
| Background noise (0) | 1,37E+04 | 1,14E+04 | 1,80E+04 | 1,16E+04 | 1,56E+04 |
| Aerosol emission at 25 cm without protection (1) | 1,06E+06 | 6,83E+05 | 1,24E+06 | 8,93E+05 | 1,08E+06 |
| Aerosol emission at 25 cm + mask on receiver (2) | 8,27E+05 | 3,87E+05 | 1,08E+06 | 6,33E+05 | 9,45E+05 |
| Aerosol emission at 25 cm + face shield on receiver (2A) | 4,68E+05 | 2,20E+05 | 8,03E+05 | 3,23E+05 | 5,52E+05 |
| Aerosol emission at 25 cm + mask on emitter (3) | 2,93E+04 | 2,16E+04 | 5,34E+04 | 2,63E+04 | 3,30E+04 |
| Aerosol emission at 25 cm + face shield on emitter (3A) | 2,36E+04 | 1,67E+04 | 3,61E+04 | 2,10E+04 | 2,93E+04 |
| Aerosol emission at 25 cm + mask on receiver + mask on emitter (4) | 2,60E+04 | 1,61E+04 | 3,94E+04 | 2,36E+04 | 3,22E+04 |
| Aerosol emission at 25 cm + face shield on receiver + face shield on emitter (4A) | 1,93E+04 | 1,51E+04 | 2,57E+04 | 1,56E+04 | 2,35E+04 |
