## Appendix Table 2 for "Experimental efficacy of the face shield and the mask against emitted and potentially received particles" (from another version of this preprint: version 1, 2020-11-24)

**Table 2:** Modelisation with each experience compared to background level of particulate pollution

| | log(n) | | |
|---|---|---|---|
| *Predictors* | *Estimates* | *CI* | *p* |
| Background noise (0) | Ref. | | |
| Aerosol emission at 25 cm without protection (1) | 5.41 | 5.18 – 5.63 | **<0.001** |
| Aerosol emission at 25 cm + mask on receiver (2) | 3.75 | 3.53 – 3.98 | **<0.001** |
| Aerosol emission at 25 cm + face shield on receiver (2A) | 2.88 | 2.65 – 3.11 | **<0.001** |
| Aerosol emission at 25 cm + mask on emitter (3) | 0.61 | 0.38 – 0.83 | **<0.001** |
| Aerosol emission at 25 cm + face shield on emitter (3A) | 0.39 | 0.17 – 0.62 | **0.001** |
| Aerosol emission at 25 cm + mask on receiver + mask on emitter (4) | 0.42 | 0.20 – 0.65 | **<0.001** |
| Aerosol emission at 25 cm + face shield on receiver + face shield on emitter (4A) | 0.24 | 0.01 – 0.46 | **0.040** |
| Particule [0,5] | -1.05 | -1.24 – -0.87 | **<0.001** |
| Particule [1] | -2.70 | -2.88 – -2.52 | **<0.001** |
| Particule [2,5] | -4.45 | -4.63 – -4.26 | **<0.001** |
| Particule [5] | -5.67 | -5.86 – -5.49 | **<0.001** |
| Particule [10] | -6.94 | -7.12 – -6.75 | **<0.001** |
