## Appendix Table 3 for "Experimental efficacy of the face shield and the mask against emitted and potentially received particles" (from another version of this preprint: version 1, 2020-11-24)

**Table 3:** Modelisation with each experience compared to aerosol emission at 25 cm without protection

| | | **log(n)** | |
|---|---|---|---|
| *Predictors* | *Estimates* | *CI* | *p* |
| (Intercept) | 14.61 | 14.41 – 14.81 | **<0.001** |
| Aerosol emission at 25 cm without protection (1) | Ref. | | |
| Background noise (0) | -5.41 | -5.63 – -5.18 | **<0.001** |
| Aerosol emission at 25 cm + mask on receiver (2) | -1.65 | -1.88 – -1.43 | **<0.001** |
| Aerosol emission at 25 cm + face shield on receiver (2A) | -2.53 | -2.75 – -2.30 | **<0.001** |
| Aerosol emission at 25 cm + mask on emitter (3) | -4.80 | -5.02 – -4.57 | **<0.001** |
| Aerosol emission at 25 cm + face shield on emitter (3A) | -5.01 | -5.24 – -4.79 | **<0.001** |
| Aerosol emission at 25 cm + mask on receiver + mask on emitter (4) | -4.98 | -5.21 – -4.76 | **<0.001** |
| Aerosol emission at 25 cm + face shield on receiver + face shield on emitter (4A) | -5.17 | -5.40 – -4.94 | **<0.001** |
| Particule [0,5$\boldsymbol{\mu}$] | -1.05 | -1.24 – -0.87 | **<0.001** |
| Particule [1$\boldsymbol{\mu}$] | -2.70 | -2.88 – -2.52 | **<0.001** |
| Particule [2,5$\boldsymbol{\mu}$] | -4.45 | -4.63 – -4.26 | **<0.001** |
| Particule [5$\boldsymbol{\mu}$] | -5.67 | -5.86 – -5.49 | **<0.001** |
| Particule [10$\boldsymbol{\mu}$] | -6.94 | -7.12 – -6.75 | **<0.001** |
