## Appendix Table 4 for "Experimental efficacy of the face shield and the mask against emitted and potentially received particles" (from another version of this preprint: version 1, 2020-11-24)

**Table 4:** Reduction factor in the different situations, for particles ≤0.3µm

| **Situations for particles ≤0.3µm** | **Mean** | **Median** | **SD** | **Reduction factor** | ***p*-value** |
|---|---|---|---|---|---|
| Background (0) | 9581 | 9349 | 1226 | | |
| No protection (1) | 600477 | 638785 | 120624 | | |
| Mask on the receiver (2) | 521304 | 563144 | 153538 | 0.132 | **0.004** |
| Face shield on the receiver (2A) | 312319 | 319466 | 118576 | 0.48 | |
| Mask on the emitter (3) | 23749 | 21980 | 7404 | 0.96 | NS |
| Face shield on the emitter (3A) | 18340 | 17360 | 4967 | 0.969 | |
| Double protection Masks (4) | 19330 | 18542 | 6041 | 0.968 | **0.0237** |
| Double protection Face shields (4A) | 13799 | 13627 | 3071 | 0.977 | |
