## Appendix Table 5 for "Experimental efficacy of the face shield and the mask against emitted and potentially received particles" (from another version of this preprint: version 1, 2020-11-24)

**Table 5:** Sensitivity analyses in microparticle subgroups

| | log(n) | | |
|---|---|---|---|
| *Predictors* | *Estimates* | *CI* | *p* |
| PPE location [Both] | -0.10 | -0.28 – 0.09 | 0.296 |
| PPE location [Receptor] | 3.16 | 2.97 – 3.34 | **<0.001** |
| Protection [Mask] | 0.38 | 0.23 – 0.53 | **<0.001** |
| Particule [0,5] | -1.06 | -1.15 – -0.97 | **<0.001** |
| Particule [1] | -2.88 | -2.96 – -2.79 | **<0.001** |
| Particule [2,5] | -4.71 | -4.79 – -4.62 | **<0.001** |
