## Appendix Table 6 for "Experimental efficacy of the face shield and the mask against emitted and potentially received particles" (from another version of this preprint: version 1, 2020-11-24)

**Table 6:** Sensitivity analyses in macroparticle subgroups

| | log(n) | | |
|---|---|---|---|
| *Predictors* | *Estimates* | *CI* | *p* |
| Location [Both] | -0.31 | -0.60 – -0.03 | **0.030** |
| Location [Receptor] | 2.14 | 1.85 – 2.42 | **<0.001** |
| Protection [Mask] | 0.52 | 0.29 – 0.75 | **<0.001** |
| Particule [5] | 1.46 | 1.34 – 1.59 | **<0.001** |
